# Bioinformatics of Potential Biomarkers in Patients with Repeated Implantation Failure

**DOI:** 10.1101/2020.03.29.20046847

**Authors:** Mingxia Gao, Yue Kang, Bin Li, Shuang Fan, Xuehong Zhang

## Abstract

**Objective:** This study applied the public genetic databases high-throughput gene expression omnibus (GEO) database to compare the difference of gene screening between the repeated implantation failure (RIF) patients and women with normal endometrium. This study aims to provide additional information for the diagnosis and treatment in RIF patients through the gene set enrichment analysis and the construction of protein–protein interactions (PPIs) network.

**Method:** The human endometrial microarray data of RIF and normal control group were obtained from the GEO database provided by the National Center for Biotechnology Information (NCBI). The analysis and the review of differential gene screening, gene ontology (GO) pathway analysis, Kyoto Encyclopedia of Genes and Genomes (KEGG) analysis, and PPIs network construction were carried out.

**Result:** 273 genes with differential expression, 87 up-regulated and 186 down-regulated genes, were obtained through differential gene analysis. 50 genes with the most significant differential expression fold change were screened for subsequent analysis. KEGG results indicated that the top three related pathways were cancer pathway, MAPK signaling pathway, and homologous recombination pathway when the relevant pathways were sequenced according to the correlation size. GO analysis showed that differentially expressed genes were involved in protein phosphorylation, complement activation, and other biological processes. PPIs analysis demonstrated that targeted genes show high correlation to the endometrial receptivity, including PAEP, CXCL14, HOXB3, CD55, and VEGFA, all were down-regulated in RIF endometrial tissues compared to the control group, and these genes also influenced and regulated each other.

**Conclusion:** Through public databases and bioinformatics, several factors in endometrial receptivity that affect RIF directly and indirectly were found. In particular, the immune and angiogenesis factors stood out the most. The different genes which regulate the immune response and angiogenesis response may become new targeted treatment for RIF.

## Introduction

With the rapid development of in vitro fertilization-embryo transfer (IVF-ET) in recent years, the quantity and quality of embryos have been optimized. However, the multiple embryo transfers were performed frequently due to relatively low pregnancy rates in IVF-ET patients^[1, 2]^, and many patients suffer from repeated implantation failure (RIF) through the procedure. Though the term is not clearly defined, RIF is deemed when the patient fails to get a clinical pregnancy after three cycles of IVF-ET or after at least four good-quality embryo transfers^[3,4,5]^. Previous studies on RIF were mostly conducted in clinical trials, and the limited amount of biomarkers were detected in the peripheral blood; few studies investigated the local endometrial tissues and RIF gene expression.

The key factors for RIF were controversial. Comparing the endometrial gene expression profiles between RIF patients and healthy women, and investigating the factors affecting endometrial receptivity and the mechanism of RIF could provide new biomarkers to the diagnosis of RIF patients as insufficient or damaged endometrial receptivity might be one of the main factors related to RIF. This new finding aims to provide additional information for the diagnosis and treatment in RIF patients.

Endometrial receptivity refers to the state of the endometrium during the implantation window (WOI), which is considered to be the most receptive period for embryo implantation, preparing a favorable environment in the endometrium^[1,9]^. Inflammatory reaction, immune response, and angiogenesis all had influence on endometrial receptivity^[10,11,12]^. However, although multiple factors were involved in the occurrence and development of endometrial receptivity, the specific molecular mechanism was still unclear^[13]^.

In this study, high throughput gene expression omnibus (GEO) was used to screen related genes in receptivity of human endometrium, and enrichment analysis of differential gene functions was conducted. Meanwhile, the PPIs network was constructed to provide more targets in diagnostic and therapeutic for patients with RIF.

## 1. Method

### 1.1 Genetic data sources and experimental groupings

The GSE103465 packet was selected (Figure 1) from GEO database (www.ncbi.nlm.nih.gov/geo/) microarray platform GPL16043 gene expression arrays (people) provided by the National Center for Biotechnology Information (NCBI) to obtain microarray data of human endometrial genes in subjects with RIF and healthy women without RIF. Gene expression data were uploaded by Guo F, Si C et al(the packet name was GSE103465), among which three patients were in the group of RIF and three were in the healthy control group. GSE103465 data has not been published.

**Figure 1.**
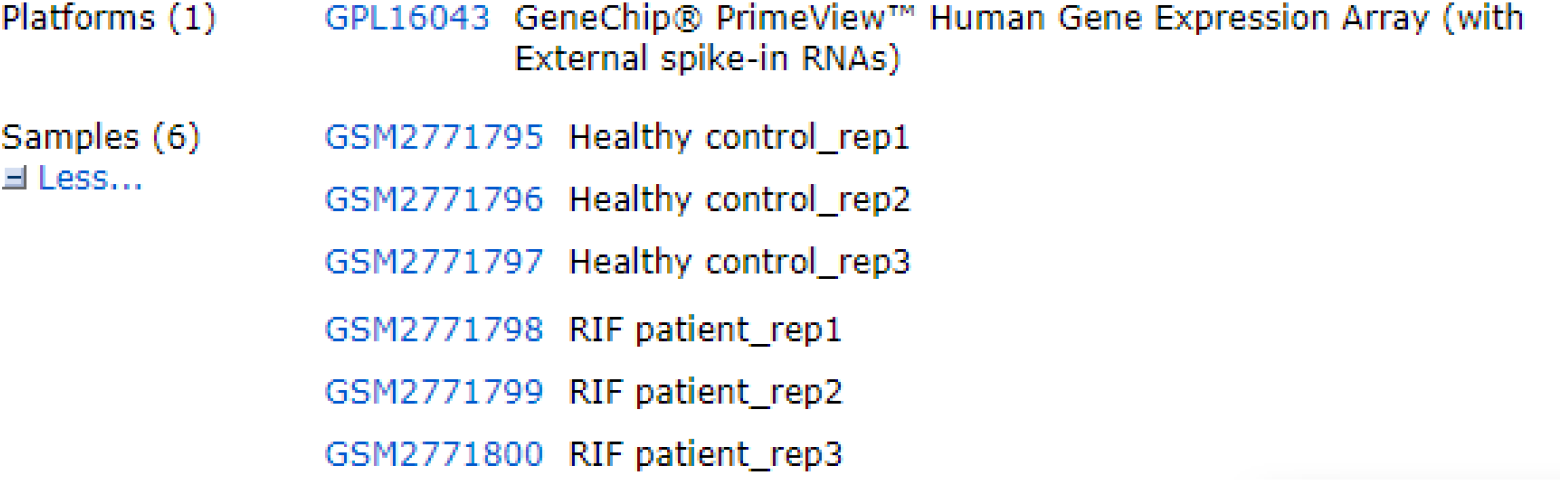

### 1.2 Differential gene screening

The original data were standardized and processed using the online tool GEO2R/R software package LIMMA (Linear Models for Microarray Analysis). In this study, p-value ≤0.05 and log|FC|≥1 were used as the cutoff values to screen for differential genes between the RIF and the healthy control group.

### 1.3 Analysis of gene ontology (GO) and kyoto encyclopedia of genes and genomes (KEGG)

Differential genes were analyzed using GO and KEGG pathways. A large-scale genomic data of the differential genes was examined through DAVID Functional Annotation Bioinformatics Microarray Analysis^[14,15]^ (https://david-d.ncifcrf.gov/) regarding their functional roles and signal pathway involvement in biological processes, cell components, and molecular functions. Annotation, visualization, and integrated analysis were performed to explore the association between the genomic data with RIF.

### 1.4 PPIs network construction

Functional protein association network (STRING, http://string-db.org/) database was used to construct the PPIs network. In this study, PPIs confidence >0.4 was the selection threshold for PPIs network construction, and the central node (the corresponding protein of the central node was the core protein with important physiological functions) was screened by calculating the edge of the node. The node color varies according to log|FC|.

## 2. Results

### 2.1 Screening in different expression of genes

Stratified clustering showed all differentially expressed genes (Figure 2). The box plot showed that after normalization, the distribution of differential genes in 6 control samples was basically the same (Figure 3A). Through the statistical analysis of MicroRNA(miRNA) expression data, the endometrium of women with RIF and the endometrium in healthy control group were identified for miRNA with differential expression. In the scatter diagram shown in Figure 3(B), the different expression of miRNAs were located on the left and right of the vertical axis of the top coordinate.

**Figure 2.**
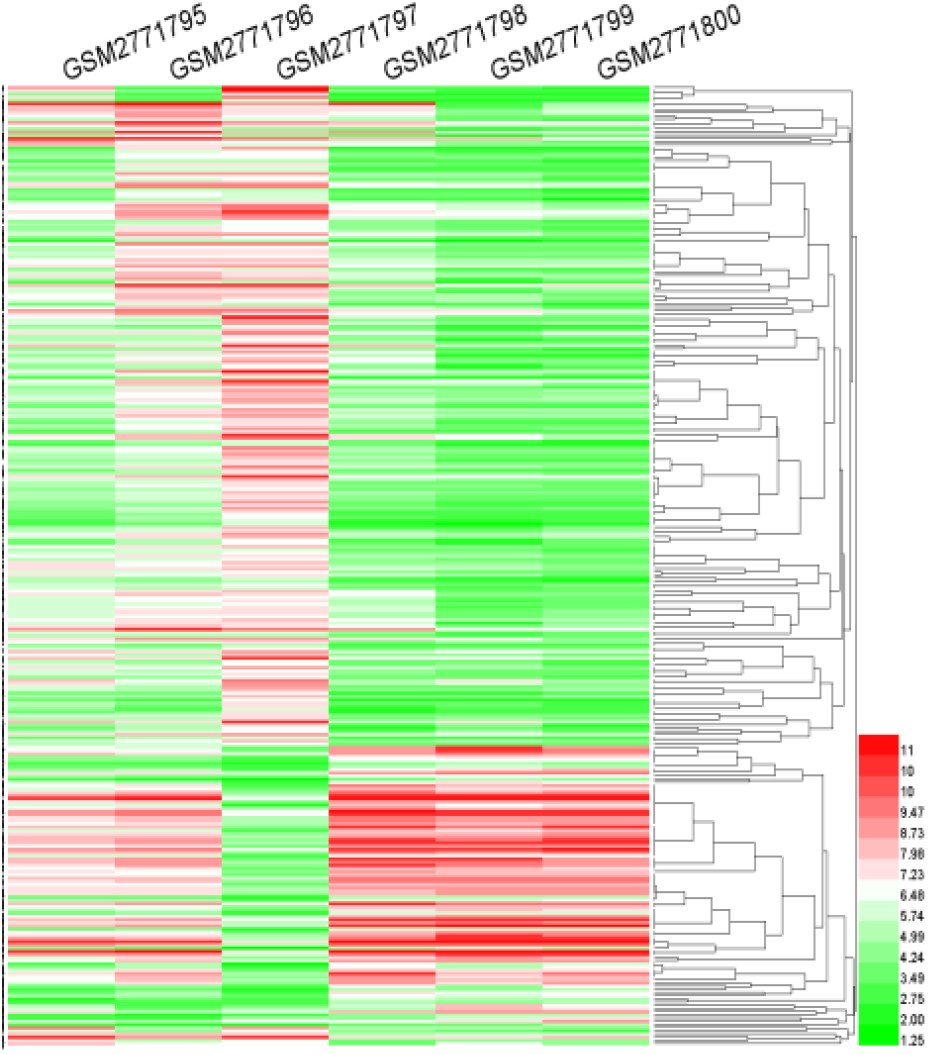
Cluster analysis revealed miRNA expression patterns with differential expression between samples. Normalization and data conversion using the R software LIMMA package through the GEO2R platform.

**Figure 3.**
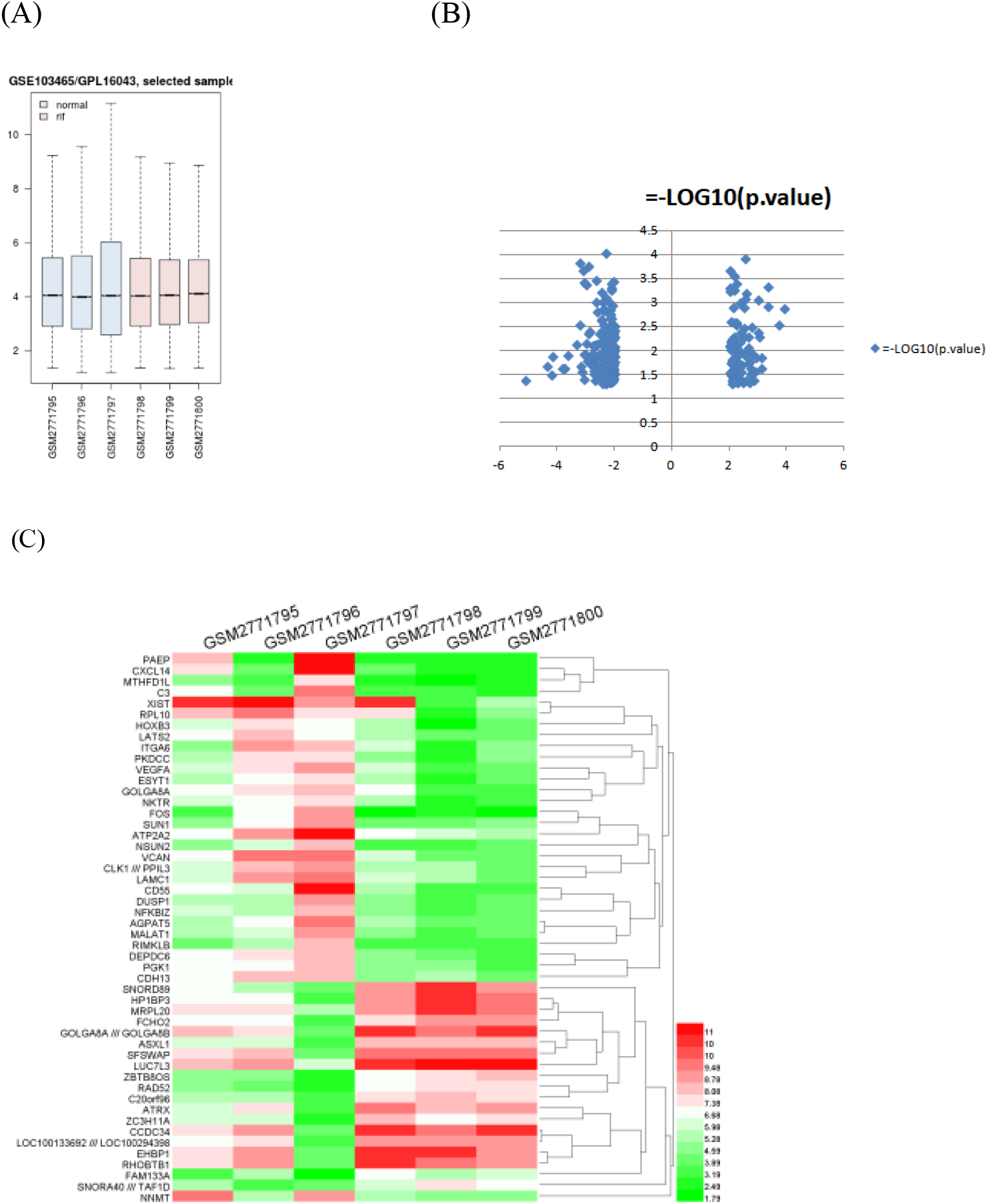
Analysis of differentially expressed miRNA in patients with RIF: (A) box plot indicates that the distributions of differentially expressed genes in 6 control samples were similar after normalization. (B) the dots on both sides of the vertical axis in the scatter plot exhibits the up-down-regulated differentially expressed genes, with changes of more than 2 times. (C) heat map shows differential expression multiples in the top 50 genes. Red indicates high relative expression; green indicates low relative expression.

The differentially expressed genes were sequenced according to their differentially expressed multiples (FC≥2.0). There were 273 differentially expressed genes, including 87 up-regulated genes and 186 down-regulated genes. A heat map was drawn to show the genes in the top 50 expression multiples of all differentially expressed genes (Figure 3C).

The GSE103465 data package was analyzed through the GEO2R platform, the LIMA package of R software was used for data normalization processing, and the basic functions of R software were used for data conversion. The differentially expressed genes between two samples were identified by differential expression multiple screening, and the gene expression patterns between different samples were revealed by hierarchical clustering method. The software package and heat map in R software were used to visualize the analysis. Red indicates up-regulation, and green indicates down-regulation.

### 2.2 Analysis of GO and KEGG

GO analysis includes three parts: molecular function (MF), biological process (BP), and cell component (CC). 60 genes with the most significant differential expression fold change were selected for GO analysis. Meanwhile, KEGG analysis of these differentially expressed genes was conducted using the DAVID database. Top 50 genes with the largest-log (*p*-value) were selected in the bar chart of KEGG, which participated in multiple signaling pathways and might play an critical role in the occurrence of some specific diseases.

GO analysis of differentially expressed genes (Figure 4A) showed that they were mainly involved in biological processes such as biological regulation, protein phosphorylation, molecular function regulation. Including the level of cell components: differentially expressed genes are mainly involved in the cell components such as the nucleotide and the Golgi apparatus. Molecular functional analysis showed that the differentially expressed genes were involved in protein kinase activity, protein phosphorylation, complement pathway activation, and some other processes.

**Figure 4.**
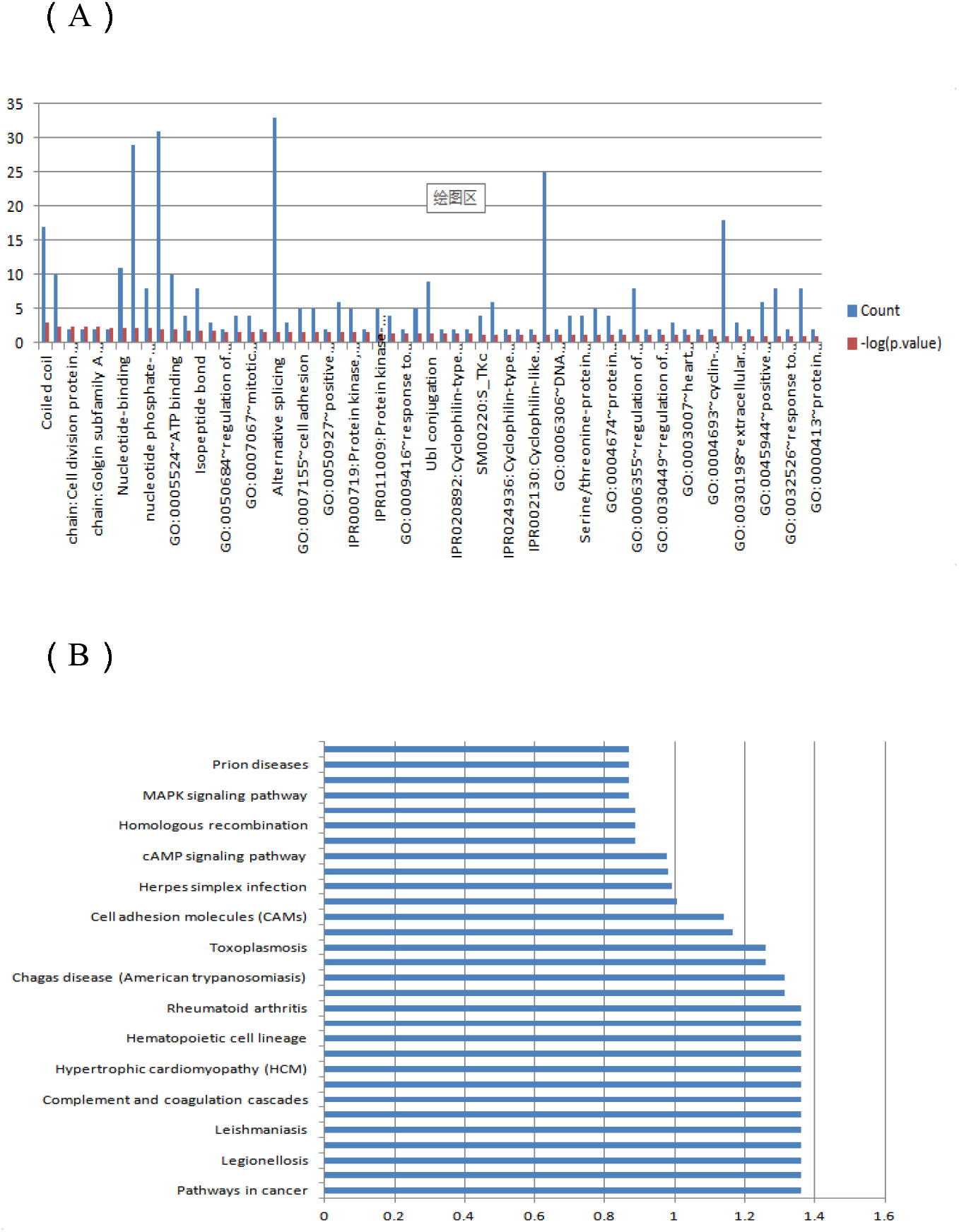
GO analysis of differentially expressed genes and notes of KEGG pathway :(A) biological process, cell composition, and molecular function analysis of differentially expressed genes. (B) KEGG pathway analysis revealed the first 15 pathways related to differentially expressed genes.

KEGG analysis (Figure. 4B) revealed 15 pathways associated with differentially expressed genes, including MAPK signaling pathways, alanine, aspartic and glutamate metabolism, cytokine receptor interactions, and prion diseases. When ranking related pathways according to their correlation size, the top three related pathways were cancer pathway, MAPK signaling pathway, and homologous recombination (Figure. 4B).

Among them, biological process analysis of differentially expressed genes (Figure 4A) found that target genes were mainly related to nucleotide modification, protein modification, macromolecular binding and other processes. Cell composition analysis (Figure. 4A) showed that the target genes were mainly related to nucleotide, Golgi body, outer envelope and other cell components. Molecular function analysis (Figure 4A) showed that differentially expressed genes were involved in protein phosphorylation, addiment activation, and other biological processes.

### 2.3 PPIs network analysis

The platform STRING (https://string-db.org/) was used to achieve the PPIs network analysis and construct the plots. There were genetic adjacency relations among vascular endothelial growth factor (VEGF), immune-related pathways represented by CD55 gene, progestogen-associated endometrial protein (PAEP), phosphoglycerate kinase 1 (PGK1), multi-function proteoglycan (VCAN), dual specificity phosphatase 1 (DUSP1), and addiment C3 (C3). Co-expressions among VEGF, C3, and DUSP1 were also revealed in the analysis.

VCAN gene belongs to aggrecan/versican proteoglycan family. The encoded protein is the main component of the extracellular matrix and plays a vital role in tissue formation and maintenance. This leads to the hypothecation that VEGFA co-regulates the production of endometrial blood vessels. The protein is involved in cell adhesion, proliferation, migration, and angiogenesis. The protein encoded by DUSP1 gene was a dual-specificity phosphatase for tyrosine and threonine, which could dephosphorylate MAP kinase MAPK1/ERK2 and participate in various cell processes. By combining with KEGG analysis, it was considered to be involved in the regulation of MAPK signaling pathway. At the same time, the PGK1 gene encoding protein was a glycolytic enzyme, catalyzing 1,3-biphosphoglycerate into glycerol 3-phosphate, the coding protein also could be a polymerase cofactors. In addition, the protein secreted by tumor cells participates in angiogenesis. It reduces the disulfide bond of the serine protease (plasmin), leading to the release of tumor blood vessels inhibin. Since PGK1 encoding protein was adjacent to VEGF gene, it was speculated that might play a certain role in regulating angiogenesis. The most noteworthy was that, as shown in figure 5, VEGF and CD55 gene has a direct adjacency relation, and the same relationship between CD55 and C3 is indicated. From the above, it showed that angiogenic pathway and complement mediated inflammatory response has a complex relationship, and they work together to determine the quality of the endometrial receptivity. To further support the previous statement, the diagram also showed that PAEP and VEGF has a genetic adjacency relations.

**Figure 5.**
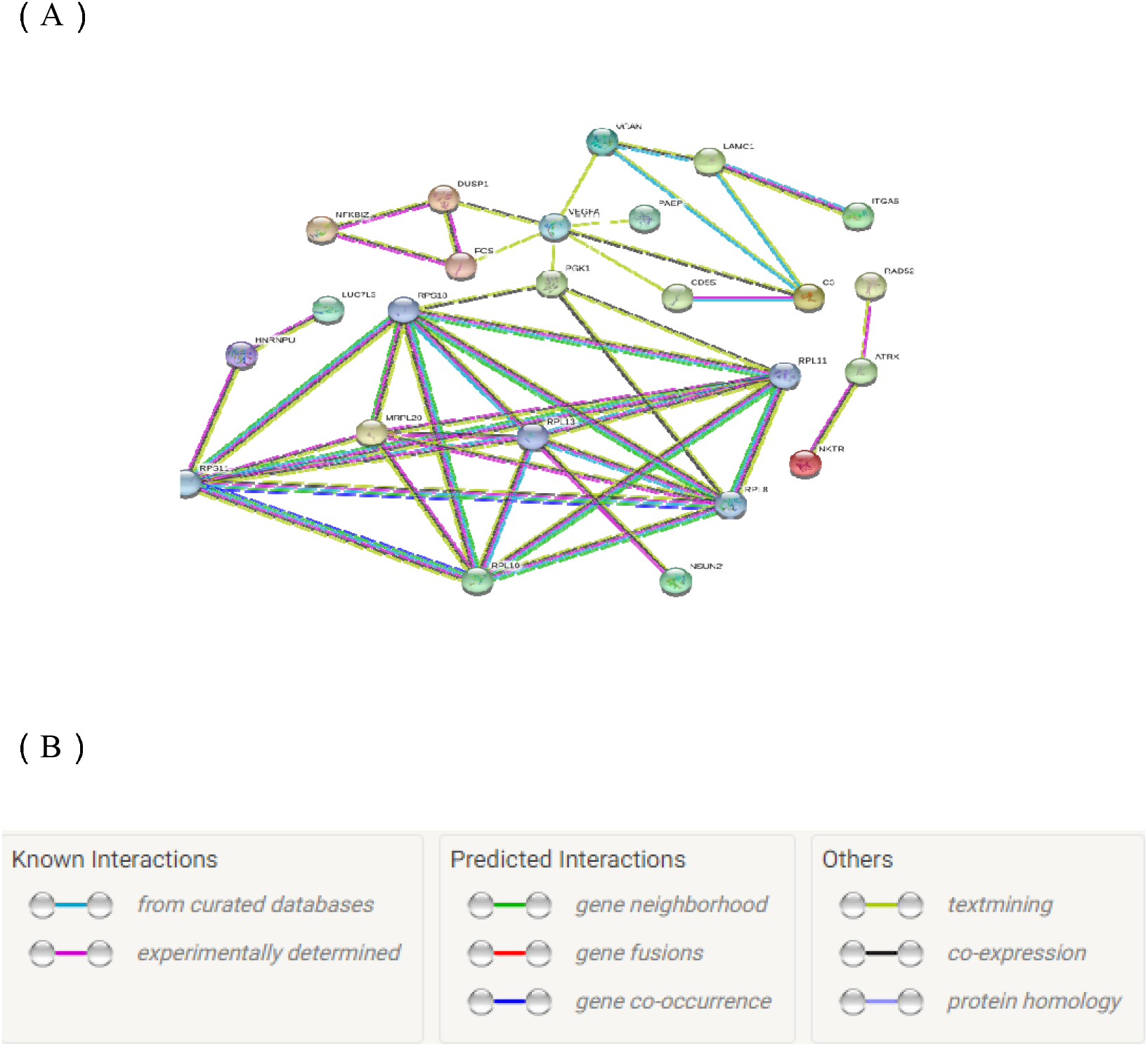

## 3. Discussion

This study applied the public genetic databases high-throughput gene expression omnibus (GEO) database to compare the difference of gene screening between the repeated implantation failure (RIF) patients and women with normal endometrium. The comprehensive human genes in IVF-ET patients that are relevant to the endometrial receptivity were analyzed through bioinformatics analysis and the differentially expressed genes screening. This study focuses on improving diagnostic method and treatment strategy for RIF patients.

49,495 genes related to human endometrial receptivity in RIF patients and healthy controls were obtained from human endometrial GSE103465 gene chip data, the GEO database microarray platform GPL16043 gene expression arrays (people) provided by the National Center for Biotechnology Information (NCBI) (www.ncbi.nlm.nih.gov/geo/). 273 genes have differences in the expression, of which 87 genes were up-regulated and 186 genes were down-regulated. The genes of interest with high correlation with the endometrial receptivity include PAEP, CXC chemokine ligand14 (CXCL14), Homeobox B3 (HOXB3), CD55, and VEGF. Among them, the expressions of PAEP, CXCL14, HOXB3, CD55, and VEGF were upregulated in normal endometrial tissues in healthy women compared with those who underwent RIF. PAEP is a protein-coding gene that encodes glycoproteins once known as pregnancy-associated endometrial alpha 2-globulin, placental protein14, and glycoprotein, and have been formally named progesterone-associated endometrial protein. The glycoproteins encoded by PAEP are mainly found in amniotic fluid, follicular fluid, and seminal plasma. The glycoproteins have the same protein framework with different glycosylation characteristics, which regulates the key steps in the fertilization process and plays an immune regulatory role. Four sugar forms of these glycoproteins: placental protein-s, -a, -f, and -c have been identified in reproductive tissues with different glycosylation and biological activities. Placental protein-a had contraceptive and immunosuppressive effects^[14,15]^. Placental protein-s could stimulate sperm to bind to zona pellucida^[16]^. Placental protein-f could inhibit sperm binding to zona pellucida and significantly inhibit progesterone-induced sperm acrosomal reaction^[17]^. Placental protein-s in seminal plasma maintained the incapacitation state of human sperm^[18]^.

CXCL14 is a protein-coding gene, which belongs to the cytokine gene family and encodes secretory proteins that are involved in immune regulation and inflammatory processes. It was also known as breast and kidney-expressed chemokine (BRAK), which was a chemokines for expressed in the breast and kidney, and it can inhibit angiogenesis^[19]^. The protein encoded by this gene was structurally related to the CXC (cys-x-cys) subfamily of cytokine, showing chemotactic activity to monocytes, which was also an enhancer and activator to dendritic cells, participating in homing of these cells^[20]^, and stimulating the migration of activated NK cells^[21]^. NCBI analysis of the gene pool suggested that CXCL14-related signaling pathways include ERK signaling pathway and PEDF induction signaling pathway.

HOXB3 is also a protein-coding gene. NCBI provided the related pathways, including the transcription and developmental biology of the androgen receptor (AR) regulatory genes KLK2 and KLK3, which encode proteins involved in hematopoietic regulation ^[22]^.

The VEGF gene, which belongs to the PDGF/VEGF growth factor family, encodes a heparin-binding protein. The protein exists in the form of disulfide bond homodimer, and induces the proliferation and migration of vascular endothelial cells, which is crucial for angiogenesis in both physiological and pathological processes. This gene encodes a kind of protein that is positively correlated with endometrial receptivity^[23]^, suggesting that the formation of blood vessels is helpful to improve endometrial receptivity.

Then, we used the online tool GEO2R LIMMA/R package for original data standardization and data processing, with p-value ≤ 0.05 and log|FC|≥1 for the boundary value, selected RIF group and the healthy control group. The top 50 places with the largest differential expression multiples were selected, then the Cluster analysis, GO and KEGG pathway analysis were performed.

KEGG analysis showed that 15 pathways were associated with differentially expressed genes in endometrium, including MAPK signaling pathways, alanine, aspartic and glutamate metabolism, cytokine receptor interactions, and prion diseases. The top three pathways were cancer pathway, MAPK signaling pathway, and homologous recombination when the associated pathways were sequenced according to the correlation size, which strongly suggested that differentially expressed genes could play an important role in cancer, protein activation, and cell division. Meanwhile, as a new finding, KEGG pathway analysis suggested that differentially expressed genes also play an important role in the complement system, MAPK signaling pathway, and angiogenesis pathway.

Successful embryo implantation requires good receptivity of endometrium ^[24]^. Boomsma C M demonstrated that cytokines played an important role in embryo implantation, trophoblast growth, and differentiation by regulating the immune and endocrine systems, suggesting that appropriate inflammatory responses promote endometrial receptivity to embryos ^[25]^. It was also reported that, to some extent, the activation of ERK1/2 -mTOR pathway could improve the endometrial receptivity during the WOI, and ERK1/2 pathway was one of the MAPK pathway ^[26]^. These results indicated that the signal pathway of differentially expressed genes was closely related to embryo implantation.

Meanwhile, GO analysis showed that differentially expressed genes in endometrium were involved in biological processes such as biological regulation, protein phosphorylation, and molecular function regulation. Differentially expressed genes were mainly involved in the nucleotide and Golgi apparatus at the cellular level. In terms of molecular function analysis, differentially expressed genes were involved in protein kinase activity, protein phosphorylation, complement pathway activation, and other processes. The analysis further demonstrated that endometrial receptivity was closely linked to the formation of blood vessels and inflammation occurrence.

An adjacency relation between VEGF and immune-related pathways represented by CD55, PAEP, PGK1, VCAN, DUSP1, PGK1, and C3 were found in PPIs analysis. It fully proved its connection to gene expression and molecular function. VEGF was also co-expressed with C3 and DUSP1, suggesting that the angiogenesis pathway was affected by biological processes such as inflammatory response and protein phosphorylation as well as the occurrence of inflammation and angiogenesis. These biological processes directly or indirectly determined the quality of the endometrial receptivity.

To sum up, this study applied enrichment analysis and pathway analysis of differential genes related to endometrial receptivity in the public gene database, found that several factors that affect endometrial receptivity directly or indirectly affect RIF, especially immune response and angiogenesis related factors. These genes, which regulate immune response and angiogenesis response, may become new targets for RIF treatment. In the follow-up studies, more attention should be paid on the biological functions and molecular mechanisms of differentially expressed genes, so as to further study in potential diagnostic biomarkers or therapeutic targets.

## Data Availability

Data were obtained from the GEO database provided by the National Center for Biotechnology Information (NCBI).

## Authors’ roles

M.X.G. and X.H.Z. accomplished the conception and design, Y.K. searched involved studies, Y.K. and B.L. accomplished the microarray data citing, differential genes screening and bioinformatics analysis. M.X.G. and S.F. reexamined these data. B.L. and Y.K. analyzed and explained data. The manuscript was written by M.X.G. and Y.K. X.H.Z. and S.F. modified the manuscript. All authors finished final approval of the version to be published.

## Acknowledgements

We gratefully thank the anonymous referees for their important and helpful comments on this manuscript. This study was supported by the Youth Research Program of the first hospital of Lanzhou University (no.ldyyyn2018-05). The authors have no competing interests to declare.

